# Real-World Clinical Performance of the Abbott Panbio with Nasopharyngeal, Throat and Saliva Swabs Among Symptomatic Individuals with COVID-19

**DOI:** 10.1101/2021.01.02.21249138

**Authors:** William Stokes, Byron M. Berenger, Danielle Portnoy, Brittney Scott, Jonas Szelewicki, Takshveer Singh, Allison A. Venner, LeeAnn Turnbull, Kanti Pabbaraju, Sandy Shokoples, Anita A. Wong, Kara Gill, Tracy Guttridge, Dustin Proctor, Jia Hu, Graham Tipples

## Abstract

**BACKGROUND:** Point of Care Testing (POCT) SARS-CoV-2 antigen tests, such as the Abbott Panbio, have great potential to help combat the COVID-19 pandemic. The Panbio is Health Canada approved for the detection of SARS-CoV-2 in symptomatic individuals within the first 7 days of COVID-19 symptom onset(s).

**METHODS:** Symptomatic adults recently diagnosed with COVID-19 in the community were recruited into the study. Paired nasopharyngeal (NP), throat, and saliva swabs were collected, with one paired swab tested immediately with the Panbio, and the other transported in universal transport media and tested using reverse-transcriptase polymerase chain reaction (RT-PCR). Positive percent agreement (PPA) was calculated. Subsequently, individuals within 7 days of symptom onset who presented to community assessment centres for SARS-CoV-2 testing had Panbio testing completed and paired with RT-PCR results from parallel NP or throat swabs.

**RESULTS:** 145 individuals were included in the study. Collection of throat and saliva was stopped early due to poor performance (throat PPA 57.7%, n=61, and saliva PPA 2.6%, n=41). NP swab PPA was 87.7% [n=145, 95% confidence interval 81.0% - 92.7%]. There were 1,641 symptomatic individuals tested by Panbio in community assessment centres, with 268/1641 (16.3%) positive for SARS-CoV-2. There were 37 false negatives, corresponding to a PPA of 86.2% [81.5% - 90.1%].

**CONCLUSIONS:** The Panbio test reliably detects most cases of SARS-CoV-2 from adults in the POCT community setting presenting within 7 days of symptom onset using nasopharyngeal swabs. Throat and saliva swabs are not reliable specimens for the Panbio.

## INTRODUCTION

The Panbio (Abbott, Ill, United States) is approved by Health Canada for the detection of SARS-CoV-2 antigen in individuals who are within the first 7 days of symptom(s) onset. The immunochromatographic assay detects SARS-CoV-2 nucleocapsid protein and is indicated only for nasopharyngeal (NP) swabs. The test should be conducted either immediately after collection, or up to 2 hours if the NP swab is placed in the Panbio extraction tube filled with extraction buffer at room temperature.^1^

Based on a study by Abbott that was conducted on 585 NP specimens collected from individuals exposed to SARS-CoV-2 or having COVID-19 symptoms within 7 days, sensitivity of the Panbio was found to be 91.4% and specificity 99.8% when compared to a reverse transcriptase real-time polymerase chain reaction (RT-PCR) reference method. Sensitivity increased to over 94% when samples with cycle threshold (Ct) >33 were excluded.^1^ There is currently a paucity of data available from external third parties on the Panbio’s performance. The few published studies have demonstrated Panbio sensitivity ranges from 72.6% - 86.5% among symptomatic individuals or exposed asymptomatic individuals.^2-4^

We sought to assess the positive percent agreement (PPA) of the Panbio by comparing its performance to RT-PCR testing among individuals in the community using two separate evaluations. The first by testing individuals with recently confirmed COVID-19 while adhering as closely as possible to manufacturer recommendations (testing of symptomatic individuals within 7 days of symptom(s) onset). The second setting was a prospective evaluation using the Panbio to diagnose COVID-19 in SARS-CoV-2 assessment/screening centres. We also tested the accuracy of the Panbio with samples taken from asymptomatic individuals at low risk for COVID-19 (i.e. no exposures), and on retrospective clinical samples previously positive for common respiratory pathogens.

## METHODS

### Testing individuals with known COVID-19

Individuals residing within the Calgary and Edmonton Health Zones of Alberta, Canada, who recently tested positive for SARS-CoV-2 at Alberta Precision Laboratories (APL; AB, Canada) and confirmed as cases by Alberta Health Services (AHS; AB, Canada) Public Health were recruited. Diagnostic testing was performed by a Health Canada approved SARS-CoV-2 assay or a lab developed RT-PCR assay (see below for details). Participants were identified by an AHS Public Health confirmed case list. Oral consent by phone was obtained to collect samples in the participant’s home. At the time of consent the symptoms of the individual were recorded (usually within 24h of collecting study swabs). Individuals under the age of 18, or in supportive or congregate living facilities were excluded. Eligible individuals who consented to the study were recruited to have two NP swab, two throat swabs, and a saliva sample collected by trained healthcare professionals. The University of Calgary Research Ethics board approved this study (REB20-444).

Healthcare workers previously trained in NP and throat swab collection performing the collection were given instructions on how to collect swabs from recruited COVID-19 infected individuals. For reference testing, the YOCON NP swab and universal transport media (UTM) (Yocon, Beijing, China) and ClassiqSwabs for throat in COPAN UTM-RT (COPAN Diagnostics, California, United States) were used.^5^ Saliva samples tested on the Panbio were compared to throat swabs sent for RT-PCR testing. For Panbio testing, the NP swab provided in the Panbio testing kits,(Abbott) was used, and the ClassiqSwab was used for throat. NP swabs were collected from separate nares. Throat swabs were collected from both sides of the oropharynx and the posterior pharyngeal wall under the uvula. Throat swabs were collected approximately one minute apart and collectors were asked to alternate the order in which throat swabs were collected. Saliva was collected by having the individual hold a ClassiqSwab (COPAN) in their mouth for approximately 30 seconds.

For each paired NP and throat swab, one was tested immediately on the Panbio cartridge for testing and the other was placed into UTM for RT-PCR testing. Throat and NP swabs in UTM collected for RT-PCR testing were stored at 4°C upon arrival at the laboratory and tested within 72 hours of collection. RT-PCR testing included an assay targeting the E-gene of SARS-CoV-2 developed within our laboratory (Public Health Laboratory, APL), and the Cobas® SARS-CoV-2 test on the Cobas 6800 instrument run according to the manufacturer’s instructions.^6^ For the E gene RT-PCR, 200ul of UTM were extracted on the MagMAX Express-96 or Kingfisher Flex (ABI) using the MagMAX-96 Viral RNA Isolation Kit (ThermoFisher) or the PurePrep Pathogen Kit (MolGen) according to manufacturer’s instructions, and eluted into a volume of 110ul.

For our lab-developed test, the samples were considered positive for SARS-CoV-2 when E-gene cycle threshold (Ct) value was <35. If the Ct was ≥35, amplification from the same eluate was repeated in duplicate and was considered positive if at least 2/3 results had a Ct <41. For the Cobas SARS-CoV-2 test, as per the manufacturer, a positive result was defined as 2/2 targets positive, or 1 or more targets were positive in duplicate. If 1/2 targets were positive and duplicate testing was negative, the result was considered indeterminate.

For discrepant results (Panbio positive, RT-PCR negative), the swabs in UTM were reextracted and retested in triplicate with the N2 assay from the US CDC 2019-Novel Coronavirus (2019-nCoV) Real-Time RT-PCR Diagnostic Panel using the UltraPlex 1-Step Toughmix (Quantabio, MA, USA) and on the Cobas 6800.^7^ PPA was calculated with Clopper-Pearson 95% confidence intervals. Statistical analysis was performed using Pearson Chi-squared for categorical variables and t-test for continuous variables using STATA (version 14.1).

### Negative samples and exclusivity panel

Two NP swabs were collected from asymptomatic individuals at low risk of having COVID-19 (no recent travel, no exposures). One swab was tested immediately on the Panbio testing cartridge. The other swab was tested by RT-PCR, as explained above. To assess for cross-reactivity, retrospective samples containing various respiratory viruses, stored in UTM at −80°C, were tested by aliquoting 3 drops of sample into the Panbio testing cartridge. These samples were previously detected by the NxTAG® Respiratory Pathogen Panel (Luminex, Tx, United States) or the CDC influenza A/B multiplex assay.^8^ The ability of the Panbio to process this volume of UTM was confirmed by testing retrospective positive SARS-CoV-2 samples, in UTM, and showing that they could be detected (data not shown; only samples with Ct <25 were detectable on the Panbio).

### Prospective testing of individuals with suspected COVID-19

After the first clinical evaluation, a pilot implementation was conducted where individuals that presented to AHS assessment centres in Edmonton and Calgary, who had symptoms at time of testing and were within the first 7 days of symptom onset, were eligible for Point of Care Testing (POCT) using the Abbott Panbio test. Individuals were asked by AHS staff if they would like to receive Panbio testing or routine testing alone. Those receiving routine testing alone were not included in the analysis. For each individual who agreed to POCT Panbio testing, one NP swab was collected for Panbio testing and one NP swab in UTM from a different nare for RT-PCR. A minority of individuals had throat swabs sent for RT-PCR due to individual refusal of a second NP swab. If an individual had a negative Panbio test, the swab sent for confirmatory testing to an APL laboratory for RT-PCR testing. Positive Panbio results were considered true positives and reported to the individual (a second swab was still sent to the lab for storage). The RT-PCR testing was performed on the APL E-gene PCR or on a Health Canada/FDA approved commercial assay. Commercial assays were site specific, and included the Allplex (Seegene, Seoul, South Korea), BDMax (Becton Dickinson, NJ, United States), Panther Fusion (Hologic, MA, United States), GeneXpert (Cepheid, CA, United States), and Simplexa (DiaSorin, Saluggia, Italy).

## RESULTS

One hundred and sixty-three individuals were recruited for the first clinical evaluation. Eighteen individuals were excluded: Three were asymptomatic at the time of COVID-19 diagnosis and at time of study recruitment, nine were symptomatic at the time of COVID-19 diagnosis but asymptomatic at time of study recruitment, four had Panbio results that were not recorded, one had the Panbio reported as negative before 15 minutes, and another was unable to be processed by RT-PCR. Individual characteristics of the remaining 145 individuals is provided in Table 1.

**Table 1:** Individual characteristics of known COVID-19 in tested with the Panbio (N=145).

| Characteristic |  |
| --- | --- |
| Male gender | 42.8% |
| Mean age in years (median, range) | 39.4 (36.0, 18.5 – 86.6) |
| Mean time from starting Panbio test to confirming positive result (median, range), N=119 (4 did not record time to positivity) | 3.8min (3, 0.5 – 15min) |
| Mean Ct value for RT-PCR positive results (median, range), N=138 | 24.7 (24.8, 15.9 – 37.9) |
| Mean duration of symptoms from collection date (median, range) | 6.1 days (6, 3 – 10 days) |
| Individuals with symptom duration $\leq 7$ days from collection date | 91.0% |

Cough was the most frequent symptom at enrollment (42.8%), followed by headache (42.1%), myalgias (41.4%), sinus congestion (36.6%), malaise (31.0%), pharyngitis (29.0%), fevers/chills (28.3%), anosmia (24.1%), ageusia (24.1%), rhinorrhea (20.0%), shortness of breath (5.5%), nausea/vomiting (3.4%), and other (17.9%), including chest pain, diarrhea, eye soreness, lymphadenopathy, loss of appetite, arthalgia, dizziness, and/or conjunctivitis. Mean duration of symptoms at the time of collection was 6.1 days (median 6, range 3 - 10 days). Ninety-one percent of individuals were within the 7-day symptom onset window. The mean E-gene Ct value for positive results from RT-PCR was 24.7 (median 24.8, range 15.9 – 37.9).

Throat and saliva sample collection was terminated early due to very poor performance, therefore only 61 and 41 individuals had a throat and saliva sample taken, respectively. There were 70.0% of throat swabs tested on the Panbio that were collected before the reference throat swab. In addition, 14.6% of swabs used for saliva testing on the Panbio were collected before both throat swabs, and the rest of the saliva swabs were tested after the two throat swabs. The PPA of throat and saliva swabs (95% confidence intervals (CI)) was 57.7% (95% CI 43.2% - 71.3%) and 2.6% (95% CI 0.06% - 13.5%), respectively (see supplementary material).

Of the 145 that underwent a NP swab, 121 were positive on both the Panbio and RT-PCR (Table 2). The PPA of the Panbio compared to RT-PCR was 87.7% (95% CI 81.0% - 92.7%) (Table 3). There were 17 false negatives on the Panbio, with 14/17 (82.4%) having a Ct > 25 by RT-PCR and 9/17 (52.9%) with Ct > 30 on RT-PCR (Supplementary material). Two samples were from individuals outside the 7-day symptom onset window. Restricting the analysis to individuals with symptom onset ≤ 7 days did not significantly change the PPA of the Panbio, which was 88.1% (95% CI 81.1% - 93.2%) for this group. Panbio positive samples had lower Ct values on RT-PCR testing than Panbio negative samples (p<0.001; Table 4). All samples (n=7) with a Ct value ≥ 33 were negative on the Panbio.

**Table 2:** Results of Panbio and RT-PCR in symptomatic known COVID-19 infected individuals (N=145).

|  |  | RT-PCR |  |
| --- | --- | --- | --- |
|  |  | Positive | Negative |
| **Panbio** | Positive | 121 | 2 |
|  | Negative | 17 | 5 |

**Table 3:** Positive percent agreement (PPA) and negative percent agreement (NPA) between Panbio and RT-PCR in symptomatic known COVID-19 infected individuals (N=145).

|  | PPA [95% CI] | NPA [95% CI] |
| --- | --- | --- |
| **Panbio** | 87.7% [81.0% - 92.7%] | 71.4% [29.0% - 96.3%] |

CI: confidence interval

**Table 4:** Characteristics between Panbio negative and Panbio positive samples in known COVID-19 infected individuals (N=145).

|  | Panbio negative (N=22) | Panbio positive (N=123) | P-value |
| --- | --- | --- | --- |
| **Mean duration of symptoms at collection (days)** | 6.2 | 6.1 | 0.636 |
| **Symptoms $\leq 7$ days at collection** | 86.4% | 91.9% | 0.405 |
| **Mean age** | 37.1 | 39.8 | 0.394 |
| **Mean Ct value** | 30.6 | 23.8 | <0.001 |
| **Male gender** | 45.8% | 42.3% | 0.748 |

When tested in triplicate using RT-PCR followed by triplicate testing by the CDC method and testing on the Cobas 6800, neither of the two RT-PCR negative samples, with paired positive Panbio samples, resolved as positive.

There were 1,641 symptomatic individuals who were tested by POCT on the Panbio in community assessment centres from December 7 – 21, 2020, with 268/1641 (16.3%) positive for SARS-CoV-2. Individual participant characteristics are provided in Table 5. There were 37 false negatives, corresponding to a PPA of 86.2% [95% CI 81.5% - 90.1%]. Of the 37 false negative samples, 23 were positive on the Allplex, 6 on the Panther, and 8 on the Cobas. Of the 23 samples positive on the Allplex, 7 had E-gene Ct<20, 4 had Ct 20-25, 3 had Ct 25-30, 5 had Ct >30, and 4 required repeat testing as only 1 of the 3 targets were initially positive (N-gene or RdRp, all with Ct > 35).

**Table 5:** Characteristics from individuals with unknown COVID-19 status tested prospectively in POCT setting at assessment centres (N=1,641).

| Characteristic |  |
| --- | --- |
| Male gender | 40.0% |
| Mean age in years (median, range) | 40.8 (39, 5 – 90) |
| Location of assessment centre | Calgary (65.8%)<br>Edmonton (34.2%) |
| Specimen used for confirmatory RT-PCR | NP swab (94.5%)<br>Throat swab (5.5%) |
| Instrument used for RT-PCR | Seegene (71.1%)<br>Cobas (20.9%)<br>LDT (6.5%)<br>Other (1.5%)* |
| Mean E-gene Ct value for RT-PCR positive results (median, range), N=72 | 22.7 (22.1, 13.2 – 33.9) |

NP: Nasopharyngeal swab
LDT: Lab developed test (see “methods”)
\*GeneXpert, Simplexa, BDMax

Twenty asymptomatic individuals at low risk of COVID-19 were tested by POCT, all of which were negative on the Panbio and RT-PCR. All 11 retrospective samples containing other respiratory viruses tested were negative. These samples were previously positive for one of either human metapneumovirus, adenovirus, parainfluenza virus type 4, other coronavirus (NL63, HKU1, NL63), enterovirus/rhinovirus, respiratory syncytial virus, influenza A H3N2, influenza A H1N1, or influenza B.

## DISCUSSION

The initial community clinical study and the evaluation post-implementation by POCT at community assessment centres demonstrated similar PPA. Overall, the PPA of the Panbio was moderate at 86.2-87.7% compared to various RT-PCR platforms for this population. This study demonstrates that, in real world settings, acceptable sensitivity is achieved for clinical use with the Panbio, but confirmatory testing of negatives is likely necessary for most populations.

While there were two instances in the initial clinical evaluation of known COVID-19 individuals where the Panbio was positive and the RT-PCR was negative, we believe these are true positives based on several reasons. Participants recruited in our study were all recently diagnosed with COVID-19; none of the samples from the asymptomatic individuals at low risk of COVID-19 gave false positive results throughout the study; and no issues with false positive results have been identified by the Panbio manufacturer or among previous Panbio publications within the literature.^2-4^

The PPA of the Panbio varies in the literature from 72.6% to 86.5% among individuals with symptoms ≤ 7 days.^2-4^ These studies varied in terms of their study design and reference standards used. All studies examined paired NP swabs tested with the Panbio and a PCR-based platform, with two of the three studies using the Allplex (Seegene, South Korea).^3,4^ One study^2^ used the VitaPCR SARS-CoV-2 (Credo diagnostics, Singapore) that has limited data available on its performance, which may potentially explain why 7 samples were Panbio positive but VitaPCR SARS-CoV-2 negative.^9^ Gremmels *et al*. performed testing on the Panbio up to 2 hours from collection, which may account for the lower sensitivity detected (72.6%).^3^ Of the studies that examined positivity rate based on symptom duration, one study found no difference in positivity rate with duration of symptom onset^3^, whereas another found higher sensitivity in individuals with symptom onset < 7 days (sensitivity 86.5%) compared to individuals with symptom onset ≥ 7 days (sensitivity 53.8%).^4^ We found no difference in Panbio sensitivity among individuals with symptoms > 7 days, but this was limited to a very small pool of samples (N=15). All studies, including ours, found decreases in Panbio sensitivity among SARS-CoV-2 samples with higher Ct values, with sensitivity dropping when Ct values are approximately > 26.

Our study contributes to the literature on the Panbio’s performance by using alternative collection methods, such as throat and saliva swabs. Unfortunately, these specimens were proven to be inferior to NP swabs and should not be used on the Panbio. Further studies are required to determine if an alternative way to test saliva on the Panbio could prove effective (e.g. direct inoculation of saliva onto the Panbio test cartridge or saliva collected in a media). We did not evaluate nasal swabs on the Panbio because the results could have been altered by collecting NP swabs also. However, previous work done by our laboratory has shown nasal swabs to be inferior to throat swabs for the detection of SARS-CoV-2, so it would be surprising if nasal swabs proved to be as effective a specimen as NP swabs for testing on the Panbio.^6^

Our study was predominately restricted to individuals within the community who had symptoms ≤ 7 days. As such, our study was unable to provide any conclusions about the Panbio performance among individuals admitted to hospital, in congregate living facilities, who are asymptomatic, and individuals with symptoms > 7 days. Our study did not evaluate the negative percent agreement of the Panbio when applied in real-world settings. The strengths of our study include the large number of COVID-19 positive individuals recruited. In addition, we included prospective data taken from real-world settings (symptomatic individuals presenting to COVID assessment centres) and found similar results, which further reinforces our study findings. We also tested asymptomatic individuals at low risk of COVID-19 (no travel, no exposure), and tested retrospective samples positive for other respiratory viruses.

## CONCLUSIONS

In conclusion, the Panbio was able to detect most SARS-CoV-2 positive samples among individuals with symptomatic COVID-19 infection. However, its performance will miss at least 10% of people with confirmed COVID-19 infection, and therefore negative results on the Panbio obtained from individuals at high risk for COVID-19 infection should be considered presumptive until confirmed with a PCR test. Individuals who had a negative Panbio test result but have ongoing suspicion for COVID-19, could consider an alternative testing solution by retesting them using the Panbio at frequent intervals. Further studies are required to determine if repeat interval testing can increase sensitivity of the Panbio in detecting SARS-CoV-2 in real world settings. Given the speed, low-complexity and acceptable performance, the Panbio test is suitable for use in the POCT setting, especially when rapid identification of positive patients is critical. As such, they will play an impactful role in combating the COVID-19 pandemic by improving testing in settings where rapid turnaround times are much needed, such as among difficult to reach populations (e.g. homeless), inpatients with suspected nosocomial infection, in high throughput COVID-19 assessment centres, and in rural areas where access to a laboratory is limited because transportation delays are significant.

## Data Availability

Available upon request

## Acknowledgments

This work was funded using internal operating funds of Alberta Precision Laboratories and Alberta Health Services. Test kits and instruments were paid for by the Public Health Agency of Canada. We thank the AHS mobile integrated health team for collecting samples in the community and Alberta Precision Laboratory staff for assistance with testing of samples and in the development and support of the POCT program.

## REFERENCES

1. Abbott. Panbio: COVID-19 Ag Rapid Test Device. 2020. [Accessed Dec 25, 2020]. Available at: https://www.who.int/diagnostics_laboratory/eual/eul_0564_032_00_panbi_covid19_ag_rapid_test_device.pdf.

2. Fenollar F, Bouam A, Ballouche M, et al. Evaluation of the Panbio Covid-19 rapid antigen detection test device for the screening of patients with Covid-19. J Clin Microbiol. 2020:JCM.02589–20.

3. Gremmels, et al. Real-life validation of the Panbio COVID-19 antigen rapid test (Abbott) in community-dwelling subjects with symptoms of potential SARS-CoV-2 infection. MedRxiv. 2020. [Accessed Dec 25, 2020]. Available from: https://www.medrxiv.org/content/10.1101/2020.10.16.20214189v1.

4. Linares M, Pérez-Tanoira R, Carrero A, et al. Panbio antigen rapid test is reliable to diagnose SARS-CoV-2 infection in the first 7 days after the onset of symptoms. J Clin Virol. 2020;133:104659.

5. Berenger BM, Conly JM, Fonseca K, et al. Saliva collected in universal transport media is an effective, simple and high-volume amenable method to detect SARS-CoV-2. Clin Microbiol Infect. 2020:S1198-743X(20)30689-3.

6. Berenger B, Fonseca K, Schneider AR, Hu J, Zelyas N. Sensitivity of nasopharyngeal, nasal and throat swab for the detection of SARS-CoV-2. 2020. MedRxiv. doi:https://doi.org/10.1101/2020.05.05.20084889.

7. Centers for Disease Control and Prevention. CDC 2019-Novel Coronavirus (2019-nCoV) Real-Time RT-PCR Diagnostic Panel: For Emergency Use Only: Instructions for Use [Accessed Nov 30, 2020]. Available from: https://www.fda.gov/media/134922/download.

8. Selvaraju S, Selvarangan R. Evaluation of three influenza A and B real-time reverse transcription-PCR assays and a new 2009 H1N1 assay for detection of influenza viruses. J Clin Microbiol. 2010;48(11):3870–5

9. Fournier PE, Zandotti C, Ninove L, et al. Contribution of VitaPCR SARS-CoV-2 to the emergency diagnosis of COVID-19. J Clin Virol. 2020;133:104682.

