## Supplementary Material for "Real-World Clinical Performance of the Abbott Panbio with Nasopharyngeal, Throat and Saliva Swabs Among Symptomatic Individuals with COVID-19"

Table 1: Results of paired throat swabs tested on the Panbio and RT-PCR in individuals with symptomatic COVID-19 infection (n=61).

|  |  | **RT-PCR** | |
| --- | --- | --- | --- |
|  |  | Positive | Negative |
| **PanBio** | Positive | 30 | 0 |
|  | Negative | 22 | 9 |

Table 2: Positive percent agreement (PPA) and negative percent agreement (NPA) of paired throat swabs tested on the Panbio and RT-PCR in individuals with symptomatic COVID-19 infection (n=61)

|  | **PPA [95% CI]** | **NPA** |
| --- | --- | --- |
| **PanBio** | 57.7% [43.2% - 71.3%] | 100% [66.4% - 100.0%] |

CI: Confidence interval.

Table 3: Results of saliva swabs tested on the Panbio compared to throat swabs tested using RT-PCR in individuals with symptomatic COVID-19 infection (n=41).

|  |  | **RT-PCR** | |
| --- | --- | --- | --- |
|  |  | Positive | Negative |
| **PanBio** | Positive | 1 | 0 |
|  | Negative | 38 | 2 |

Table 4: Positive percent agreement (PPA) and negative percent agreement (NPA) of saliva swabs tested on the Panbio compared to throat swabs tested using RT-PCR in individuals with symptomatic COVID-19 infection (N=41).

|  | **PPA [95% CI]** | **NPA** |
| --- | --- | --- |
| **PanBio** | 2.6% [0.06% - 13.5%] | 100% [15.8% - 100%] |

CI: Confidence interval.

Table 5: Details on the Panbio negative, RT-PCR positive results (N=17). Two samples were from individuals outside the 7-day symptom onset.

| **Number** | **Ct value from RT-PCR** | **Duration of symptoms at time of collection (days)** | **Still experiencing symptoms at time of collection** | **Symptoms** |
| --- | --- | --- | --- | --- |
| 1 | 37.86 | 9 | Yes | Nasal congestion, myalgia |
| 2 | 32.79 | 5 | Yes | Nasal congestion, anosmia, rhinorrhea, malaise |
| 3 | 36.56 | 6 | Yes | Nasal congestion, rhinorrhea, pharyngitis, malaise |
| 4 | 29.21 | 6 | Yes | Cough, rhinorrhea, nasal congestion, headaches |
| 5 | 33.58 | 6 | Yes | Cough, pharyngitis, malaise |
| 6 | 21.51 | 6 | Yes | Headache, nasal congestion |
| 7 | 36.9 | 8 | Yes | Headache, nasal congestion, anosmia, ageusia, myalgia |
| 8 | 32.01 | 5 | Yes | Rhinnorhea, nasal congestion, diarrhea, myalgias, malaise |
| 9 | 27.01 | 7 | Yes | Fever, cough, headache, photophobia |
| 10 | 37.86 | 6 | Yes | Malaise, myalgias, fevers/chills |
| 11 | 27.42 | 7 | Yes | Myalgias, anosmia, ageusia |
| 12 | 24.83 | 7 | Yes | Headache, chills |
| 13 | 36.46 | 6 | Yes | Fevers/chills, myalgia, shortness of breath |
| 14 | 33.75 | 6 | Yes | Cough, rhinorrhea |
| 15 | 21.27 | 5 | Yes | Rhinorrhea, myalgia, headache |
| 16 | 26.03 | 4 | Yes | Anosmia, ageusia, nasal congestion, lymphadenopathy, chest tightness |
| 17 | 29.5 | 7 | Yes | Nasal congestion, headache, myalgia, malaise, myalgia |
| MEAN (median, range) | 30.9 (32.0, 21.3 – 37.9) | 6.2 (6, 4 – 9) | N/A | N/A |
